# Cost components of school-based oral health-promoting programs: a systematic review protocol

**DOI:** 10.1101/2023.06.02.23290900

**Authors:** Bárbara da Silva Mourthé Matoso, Viviane Elisângela Gomes, Wagner Marcenes, Kênya Valéria Micaela de Souza Noronha, Camilla Aparecida Silva de Oliveira Lima, Raquel Conceição Ferreira

**Affiliations:** Faculdade de Odontologia, Universidade Federal de Minas Gerais, Belo Horizonte, Minas Gerais, Brazil; Affordable Health Initiative, London, United Kingdom; Faculdade de Ciências Econômicas, Universidade Federal de Minas Gerais, Belo Horizonte, Minas Gerais, Brazil

**Author notes:** Corresponding author (VEG). BSMM, VEG, WM, KVMSN, CASOL, RCF, and EFF contributed to the conceptualization of the study. BSMM, VEG, WM, KVMSN, CASOL, and RCF contributed to defining the methodology of the study. BSMM, VEG, CASOL, and RCF contributed to writing the original draft. BSMM, VEG, WM, KVMSN, CASOL, and RCF contributed to writing, reviewing, and editing the final version of the manuscript.

**Keywords:** Systematic review, costs and cost analysis, dental economics, oral health, health promotion, school dentistry

## Abstract

**Introduction:** Oral health-promoting school programs play a crucial role in achieving universal coverage of oral health care by addressing oral diseases and promoting the well-being and quality of life of children and adolescents. However, a lack of studies has evaluated the costs associated with implementing these programs, which hinders decision-makers in adopting them on a large scale. This review aims to assess the cost components involved in school-based oral health-promoting programs.

**Methods:** This review will include studies that have conducted either partial or full economic evaluations, focusing on describing the cost components of oral health-promoting programs implemented in primary schools involving students aged 6 to 14. A systematic search was conducted across multiple databases: MEDLINE, The Cochrane Library, the Virtual Health Library, the NHS Economic Evaluation Database, Web of Science, Scopus, and EMBASE. Additionally, gray literature was searched using the Health Technology Assessment Database. Two independent reviewers will screen the titles and abstracts, followed by a full-text review based on predefined inclusion criteria. Data extraction and critical appraisal evaluation will also be carried out independently by two reviewers. In case of disagreements, the reviewer team will resolve them through discussion.

**Discussion:** The systematic review resulting from this protocol aims to provide evidence regarding the cost components and necessary resources for implementing and maintaining oral health-promoting school programs. This information can assist decision-makers in adopting these programs on a larger scale and effectively addressing oral diseases among children and adolescents.

**Protocol registration:** CRD42022363743.

## Introduction

Oral diseases are prevalent noncommunicable diseases worldwide, affecting approximately 3.5 billion individuals. If left untreated, these diseases can cause pain and suffering, impacting human health and well-being. To address this problem, important decisions, and actions have been taken, such as the historic resolution on oral health at the World Health Assembly, in 2021, the development and adoption of the Global Strategy on Oral Health, and the Global Oral Health Action Plan. These initiatives aim to achieve universal coverage of oral health services by 2030 [1].

School-based oral health-promoting programs play a crucial role in combating oral diseases and achieving universal coverage of oral health care. Globally, more than 90% of children in the primary school age group are enrolled in school, spending a significant portion of their time there [2]. Furthermore, schools can serve as the only and primary access point for oral health services for children and adolescents, making oral health programs implemented in schools valuable for health promotion and equality [3,4].

Oral health promotion can be effectively integrated into the school curriculum and activities, serving as a foundation for developing Health Promoting Schools (HPS) [5]. HPS is recognized as a strategic approach for promoting positive development and healthy behaviors among students [2]. Integrating oral health promotion activities into schools benefits the participating children and extends to their siblings, parents, and the community, as they gain new knowledge and adopt healthier behaviors [6].

However, there is a lack of studies evaluating the costs, cost components, and necessary resources associated with these school-based oral health-promoting programs, hindering the decision-making process for large-scale adoption of such programs [7,8]. Cost analyses, although scarce in this context, are essential for implementing evidence-based practices [9] and supporting decision-making [10]. Therefore, conducting precise and detailed cost studies to assess the resources needed for implementing and maintaining school-based oral health-promoting programs is necessary.

This systematic review aims to address this knowledge gap and is being conducted as part of the planning for assessing the implementation costs of o health care proposed by the Affordable Health Initiative model of the Health Promoting School program (https://www.affordablehealthinitiative.com/resources-dental-health-care). The Affordable Health Initiative (AHI) is a Charitable Incorporated Organization in the United Kingdom that offers a simple, scalable, and sustainable operational model for the World Health Organization’s HPS initiative. The AHI HPS model recognizes and addresses the socio-economic and socio-psychological barriers to a healthy lifestyle, focusing efforts on hygiene, diet, physical activities, tobacco and drug use, and alcohol consumption, all of which are associated with several health outcomes [11].

### Review question

What are the cost components of school-based oral health-promoting programs?

## Methods

This protocol follows the guidelines of the Preferred Reporting Items for Systematic Reviews and Meta-Analyses Protocol (PRISMA-P) [12] S1 file. The protocol has been registered in the PROSPERO database (CRD42022363743).

### Eligibility criteria

The research question and eligibility criteria were developed using the PICOS acronym (Population, Intervention, Comparator, Outcome, Study type).

### Population

This review includes studies involving students aged 6 to 14 years. Studies focusing on students under 5 and over 14 years of age will be excluded. The emphasis on elementary school is due to its widespread coverage among children and adolescents worldwide, as well as its alignment with the target population of the HPS initiative and the AHI HPS model. In Brazil, elementary education lasts 9 years and caters to students between 6 and 14 years old [13].

### Intervention and comparator

The review will consider studies that evaluated school-based oral health-promoting programs without any applicable comparator. We considered oral health-promoting school programs as the interventions aligned with the HPS initiative, aiming to create a school environment conducive to oral health, reduce risk factors for oral diseases, and improve knowledge and attitudes related to oral health [5].

Studies focusing on discontinued dental procedures within school-based oral health-promoting programs will be excluded (e.g., topical application of 4% NAF-PO_4_).

### Context

This review focuses on assessing costs related to school-based oral health-promoting programs in primary schools. Studies conducted in colleges, universities, kindergartens, or nursery schools will be excluded. The primary school level is the main focus due to its extensive coverage among children and adolescents globally, representing the target population for the HPS initiative.

### Outcomes

This review will include studies that evaluated the costs of school-based oral health-promoting programs, with a specific requirement for a detailed breakdown or disaggregation of costs, w, including a description of different cost components considered based on the analytical perspective and time horizon adopted.

### Study types

This review will include both partial or full economic evaluations that provide cost estimations along with descriptions of cost components. Model-based evaluations will be excluded unless they exclusively utilize empirical data (primary data on resource consumption and health outcome).

There will be no restrictions on language, location, or publication date. Studies will be excluded if they are letters, brief commentaries, reviews, and conference abstracts. Articles unaccessed or without full text available will also be excluded.

Additionally, articles without accessible full texts or unavailability of full texts will be excluded.

### Search strategy

The search strategy was developed, refined, and piloted with the collaboration of an experienced librarian. The exact search terms used in all databases are described in S2 file.

### Information sources

The search will be carried out in MEDLINE (via Pubmed), The Cochrane Library, Virtual Health Library, Economic Evaluation Database (via NHS CRD), Web of Science, Scopus, and EMBASE (via CAPES Portal). Grey literature will be searched in the Health Technology Assessment Database (via NHS CRD). Additionally, a manual search will be conducted on the references of the included articles to identify additional eligible studies.

### Study selection

Identified citations will be imported into EndNote® Web to remove duplicates. The remaining documents will be screened by independent reviewers using the Rayyan® blinding tool. Two blinded reviewers will assess the titles and abstracts against the inclusion criteria in the first round. Disagreements will be resolved through discussion, involving a third reviewer if necessary. The second round will involve the same process, this time accessing the full text. A PRISMA flowchart [14] will be presented to illustrate the study selection process.

### Assessment of methodological quality

Methodological quality and risk of bias in the included studies will be assessed using the quality assessment tool proposed by Andrade and colleagues [15] (S3). This tool is deemed appropriate for both full and partial economic evaluations. Two independent reviewers will complete the checklists, and disagreements will be resolved by a third reviewer. Study authors will be contacted to request missing or additional data, if necessary.

Regardless of methodological quality, all studies will undergo data extraction and synthesis. The results of the quality assessments will be presented in a tabular format, and the implications of study quality on the comprehensiveness and results of the systematic review will be discussed.

### Data collection

Two independent researchers will collect data using an adapted version of JBI Data Extraction Form for Economic Evaluation (S4). Disagreements will be resolved by a third researcher. Data extraction will comprise general and clinical data such as author, location, intervention, comparator, population, study design, data source, clinical outcome, and economic data (time horizon, analytical perspective, data source, data collection method, cost components, currency, adjustments and discounts, treatment of uncertainty, cost results and, whenever possible, the author’s conclusions regarding factors that promote or impede the cost and effects of the intervention).

### Data synthesis

The extracted data will be presented through narrative synthesis to address the objective and question of the review, providing a summary of findings and comparing results among studies, including the critical appraisal. Disaggregated presentation of results will be preferred. Relevant information such as analytical perspective, cost components, minimum and maximum value per cost component, and estimation method will be presented. Different currencies will be converted to USD using the same reference year.

## Discussion

The systematic review, derived from the proposed protocol, aims to generate evidence on the cost components and required resources for the implementation and sustenance of school-based oral health-promoting programs. It emphasizes the significance of oral health interventions as a foundational element for successfully implementing the Health Promoting School (HPS) initiative in educational institutions. This evidence can potentially guide decision-makers in adopting the AHI HPS model on a broader scale in various countries and regions where its implementation has taken place. Ultimately, this widespread adoption can lead to improved access to primary oral healthcare for a larger number of children and adolescents, thereby enhancing their oral health outcomes.

## Data Availability

All relevant data from this study will be made available upon study completion.

## Acknowledgments

The authors would like to thank Dr. Fernanda Gomes Almeida, librarian at Universidade Federal de Minas Gerais, for her contribution, and pay posthumous tribute to Prof. Efigênia Ferreira e Ferreira, who integrated the review team and led the conceptualization of the study.

## Supporting information

S1 Checklist. PRISMA-P (Preferred Reporting Items for Systematic Review and Meta-Analysis Protocols) 2015 checklist: Recommended items to address in a systematic review protocol.

S2 File. Search terms and strategy.

S3 File. Quality assessment form.

S4 File. Data extraction form.

